# Neuroimmune pleiotropy links COVID-19 outcomes to brain structural and functional imaging-derived phenotypes

**DOI:** 10.1101/2025.11.03.25339411

**Authors:** Qianyu Chen, Jun He, Brenda Cabrera-Mendoza, Ziang Xu, Dan Qiu, Renato Polimanti

**Author notes:** Correspondence: Renato Polimanti, PhD. Department of Psychiatry, Yale University School of Medicine. 60 Temple, Suite 7A, New Haven, CT 06510, USA.

## Abstract

This study investigated the pleiotropy and the underlying biology linking COVID-19 outcomes (infection, hospitalization, and severe symptoms) to brain structure and function, leveraging genome-wide data from the COVID-19 Host Genetics Initiative (up to 122,616 cases and 2,475,240 controls) and 3,935 brain imaging-derived phenotypes (IDP) from the UK Biobank (up to 33,224 participants). COVID-19 outcomes were genetically correlated with 11 IDPs related to brain connectivity, cortical structures, and white matter. With respect to these, we also identified local genetic correlations within five loci mapping genes involved in inflammatory response, immune regulation, brain development, and neuropsychiatric disorders. We also identified 305 Bonferroni-significant gene ontologies (GO), highlighting the pleiotropy linking COVID-19 to brain structure and function through developmental processes (e.g., immune system development, leukocyte differentiation, and neurogenesis) and intracellular signaling (e.g., synaptic signaling and immune response regulation). Our drug-repurposing analysis uncovered 41 compounds, including approved drugs that could potentially influence both COVID-19 outcomes and brain-related disorders. For instance, chlorpromazine is an antipsychotic that appears to also have antiviral and immunomodulatory activity. Overall, the present findings contributed to dissecting the biological mechanisms shared between COVID-19 and brain structure and function, highlighting the systemic nature of their relationship.

## INTRODUCTION

Growing evidence supports COVID-19 impact on brain health through multiple mechanisms, including inflammatory responses, vascular dysfunction, and immune activation ^1^. Neurological symptoms such as cognitive impairment, fatigue, and anosmia are frequently observed among individuals recovering from SARS-CoV-2 infection and a recent study highlighted their potential relationship with structural and functional brain alterations ^2^. Multiple neuroimaging studies of COVID-19 reported changes across multiple brain regions, including reductions in global brain size and grey matter volume and thickness, alterations in neural activities and connectivity patterns, and disruptions in regions associated with sensory processing, cognition, memory, and attention ^3–6^. Genetically informed analyses also identified pleiotropic mechanisms linking COVID-19 to changes in brain structures ^7–9^.

While previous studies contributed to disentangle the dynamics by which COVID-19 leads to neuropsychiatric symptoms, they have primarily focused on structural changes in cortical brain regions. In recent years, the advancements of magnetic resonance imaging (MRI) permitted investigators to generate an unprecedented amount of information regarding brain structure and function variation across thousands of participants. One notable effort is the UK Biobank (UKB), which is conducting a multi-modal brain imaging study scanning 100,000 individuals ^10^.

Applying a validated pipeline to perform quality control and uniformly process UKB data ^11^, it is possible to derive >3,900 brain imaging-derived phenotypes (IDP) using UKB data generated from T1-weighted and T2-weighted fluid-attenuated inversion recovery (T2-FLAIR) structural images, susceptibility-weighted MRI (swMRI), diffusion MRI (dMRI), task functional MRI (tfMRI), and resting-state functional MRI (rfMRI). Genome-wide association studies (GWAS) of UKB brain IDPs demonstrated that the genetics of brain structure and function strongly overlap with the predisposition to neuropsychiatric disorders and symptoms ^12, 13^. Additionally, genome-wide association statistics generated by these studies enabled conduct brain-wide causal inference analyses that identified potential direct effects of brain IDPs on mental health and other comorbid conditions ^14–16^. With respect to COVID-19, multimodal brain-wide phenotyping is expected to provide important insights into the dynamics of the neuropsychiatric symptoms observed in certain patients ^17^.

In the present study, we leveraged large-scale genomic datasets available from UKB ^13^ and COVID-19 Host Genetics Initiative (HGI) ^18, 19^ to assess the pleiotropic mechanisms linking COVID-19 outcomes (infection, hospitalization, and severe symptoms) to changes in regional and tissue volumes, cortical areas, cortical thickness, regional and tissue intensities, cortical grey-white contrast, white matter hyperintensity volumes, regional T2*, white matter tracts, tfMRI activation, rfMRI node amplitude, and rfMRI connectivity. By examining polygenic overlap, individual loci, shared molecular pathways, and potential drug targets, this study aimed to expand the characterization of COVID-19 impact on brain health and inform future research into the long-term neuropsychiatric consequences of COVID-19.

## SUBJECTS AND METHODS

### Data Sources

The present study was conducted leveraging genome-wide association statistics. Owing to the use of previously collected, deidentified data, the analyses conducted did not require institutional review board approval. Because no large-scale GWAS of brain IDPs is available for non-European descent populations, the analyses were limited to data generated from European-ancestry individuals.

COVID-19 genome-wide association statistics were obtained from the COVID-19 HGI ^18^. Specifically, we analyzed COVID-19 HGI Release 7 ^19^, which investigated up to 219,692 cases and over 3 million controls across 82 studies from 35 countries. In our study, we focused on three COVID-19 outcomes: i) severe COVID-19 respiratory symptoms (i.e., COVID-19 cases requiring respiratory support or resulting in death; 13,769 cases and 1,072,442 controls of European ancestry); ii) COVID-19 hospitalization (i.e., COVID-19 cases hospitalized due to infection-related symptoms; 32,519 cases and 2,062,805 controls of European ancestry); and iii) SARS-CoV-2 infection (confirmed COVID-19 cases irrespective of symptom severity (122,616 cases and 2,475,240 controls of European ancestry). HGI Covid-19 data are available at https://www.covid19hg.org/results/r7/.

Genome-wide association statistics of 3,935 brain IDPs (Supplemental Table 1) were obtained from the Oxford Brain Imaging Genetics Server ^12, 13^. These data were generated from 33,224 participants of European descent enrolled in the UKB, a population-based cohort ^20^. UKB brain IDPs consist of measures of brain structure and function, including regional volumes, cortical thickness, and functional connectivity, derived from MRI data acquired using standardized protocols to ensure consistency and reliability across participants ^10^. UKB brain IDP GWAS data are available at https://open.win.ox.ac.uk/ukbiobank/big40/.

### Global and Local Genetic Correlation Analyses

Linkage Disequilibrium Score Regression (LDSC) was used to estimate global genetic correlations (rg; i.e., shared genetic effects across the genome) among COVID-19 phenotypes and brain IDPs ^21^. LD reference panel was based on pre-computed data from European-descent populations available from 1000 Genomes Project Phase 3 ^22^. In accordance with LDSC guidelines (available at https://github.com/bulik/ldsc), GWAS datasets were processed using munge_sumstats.py and the LDSC analyses were limited to HapMap3 common variants ^23^. To maximize our discovery power, we considered as significant brain IDPs those with rg p<0.01 with at least two of the three COVID-19 outcomes (i.e., severe COVID-19 respiratory symptoms, COVID-19 hospitalization, and SARS-CoV-2 infection).

Local Analysis of [co]Variant Association (LAVA) was conducted to identify the genomic regions with shared genetic effects (local genetic correlation) between COVID-19 outcomes and brain IDPs ^24^. European-descent populations available from 1000 Genomes Project Phase 3 were used as LD reference panel. Following LAVA instructions (available at https://github.com/josefin-werme/LAVA), the analysis was conducted with respect to 2,495 quasi-independent blocks, each around 1 Mb in size. False discovery rate correction (FDR q < 0.05) was employed to account for the number of genomic regions tested. With respect to each FDR-significant genomic region, we used the UCSC Genome Browser (Kent et al., 2002) to annotate the corresponding coding genes.

Both LDSC and LAVA methods can correct for the potential biases due to sample overlap among the GWAS datasets investigated.

### Gene-Set Enrichment Analysis

We performed a gene-set enrichment analysis (GSA) using MiXeR approach ^25^ to investigate the pleiotropic mechanisms linking COVID-19 phenotypes to brain IDPs identified in the genetic correlation analysis. Specifically, GSA-MiXeR permitted us to model gene-set heritability enrichments while accounting for LD structure. GSA-MiXeR code is available at https://github.com/precimed/gsa-mixer. This analysis was restricted to gene ontologies (GO) related to biological processes, molecular functions, and cellular components. To account for multiple testing, Bonferroni correction was applied considering the number of GOs tested (p<6.31×10^-7^). To reduce GO redundancy and identify representative domains, we used REVIGO ^26^, considering a similarity threshold=0.7 and UniProt as the reference database.

### Drug-Repurposing Analysis

We conducted a drug repurposing analysis to identify molecular compounds associated with transcriptomic profiles overlapping those associated with the GOs identified by GSA-MiXeR. Specifically, we applied the gene2drug approach ^27^ to test GOs shared between COVID-19 outcomes and brain IDPs with respect to the connectivity map drug database (1.5M gene expression profiles from ∼5,000 small-molecule compounds, and ∼3,000 genetic reagents, tested in multiple cell types)^28^. FDR correction (FDR q<0.05) was applied to define statistically significant gene2drug results after accounting for the number of molecular compounds tested.

## RESULTS

### Global and Local Genetic Correlation

We identified 11 brain IDPs with significant genetic correlation (p<0.01) with at least two COVID-19 phenotypes (Figure 1; Supplemental Table 2). Among them, IDP 2834 (rfMRI connectivity ICA100 edge 406; Figure 2) showed significant genetic correlation with the three COVID-19 outcomes tested: severe COVID-19 respiratory symptoms rg=0.26; COVID-19 hospitalization rg=0.30; and SARS-CoV-2 infection rg=0.29. Two other rfMRI IDPs were also observed: rfMRI amplitudes ICA100 node 32 (IDP 2195; Supplemental Figure 1) genetically correlated with both severe COVID-19 respiratory symptoms (rg=0.28) and SARS-CoV-2 infection (rg=0.35), and rfMRI connectivity ICA100 edge 403 (IDP 2831; Supplemental Figure 2) genetically correlated with both severe COVID-19 respiratory symptoms (rg=-0.17) and COVID-19 hospitalization (rg=-0.17). Four IDPs related to parahippocampal and entorhinal cortical thickness (IDP: 1069, 1115, 1160, and 1274) were genetically correlated with severe COVID-19 respiratory symptoms (rg from 0.16 to 0.22) and COVID-19 hospitalization (rg from 0.21 to 0.26). Another IDP genetically correlated with these COVID-19 outcomes was related to the Lat-Fis-ant-Horizont area (IDP 910, rg=-0.27 – -0.26) and the mean orientation dispersion index in the medial lemniscus (IDP 1985, rg=0.18 – 0.17). Finally, two IDPs related to the cortical areas of the pars opercularis (IDPs 766 and 957) were genetically correlated with COVID-19 hospitalization (rg=-0.19 – -0.22) and SARS-CoV-2 infection (rg=-0.22 – -0.28).

**Figure 1:**
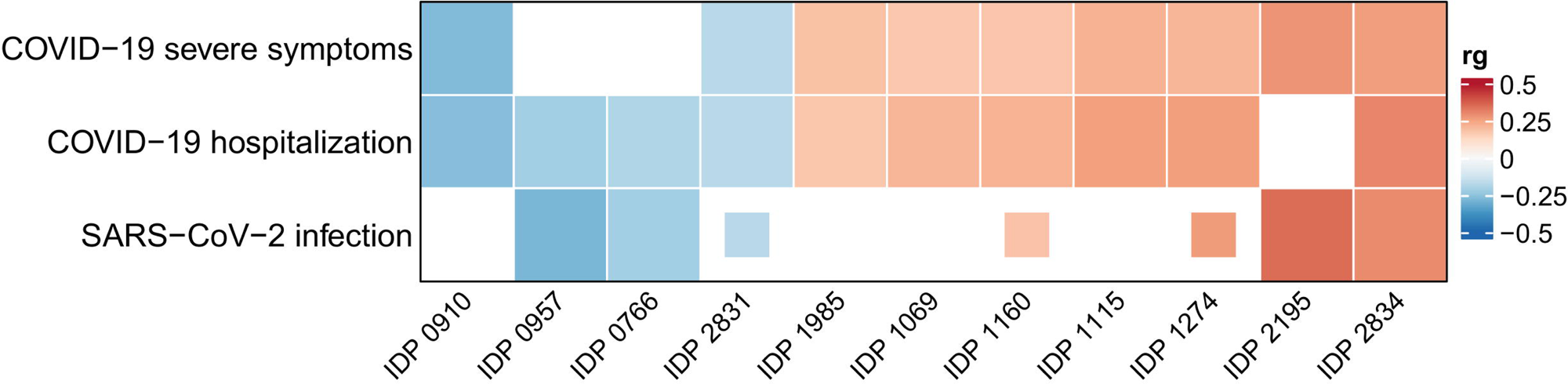
Genetic correlations between COVID-19 outcomes and brain imaging-derived phenotypes (IDP). The results reported are related to brain IDPs genetically correlated with COVID-19 outcomes at p<0.01 (fully colored cells). Nominally significant genetic correlations (0.01≤p<0.05) are indicated with partially colored cells. Blank cells represent non-significant estimates (p≥0.05). Full statistics are available in Supplemental Table 2. IDP 766: Area of parsopercularis in the right hemisphere generated by parcellation of the pial surface using Desikan-Killiany parcellation; IDP 910: Area of Lat-Fis-ant-Horizont in the left hemisphere generated by parcellation of the white surface using Destrieux (a2009s) parcellation; IDP 957: Area of G-front-inf-Opercular in the right hemisphere generated by parcellation of the white surface using Destrieux (a2009s) parcellation; 1069: Mean thickness of parahippocampal in the right hemisphere generated by parcellation of the white surface using Desikan-Killiany parcellation; IDP 1115: Mean thickness of entorhinal in the right hemisphere generated by parcellation of the white surface using BA_exvivo parcellation; IDP 1160: Mean thickness of parahippocampal in the right hemisphere generated by parcellation of the white surface using DKT parcellation; IDP 1274: Mean thickness of G-oc-temp-med-Parahip in the right hemisphere generated by parcellation of the white surface using Destrieux (a2009s) parcellation; IDP 1985: Mean OD (orientation dispersion index) in medial lemniscus (right) on FA (fractional anisotropy) skeleton (from dMRI data); IDP 2195: rfMRI amplitudes (ICA100 node 32); IDP 2831: rfMRI connectivity (ICA100 edge 403); IDP 2834: rfMRI connectivity (ICA100 edge 406).

**Figure 2:**
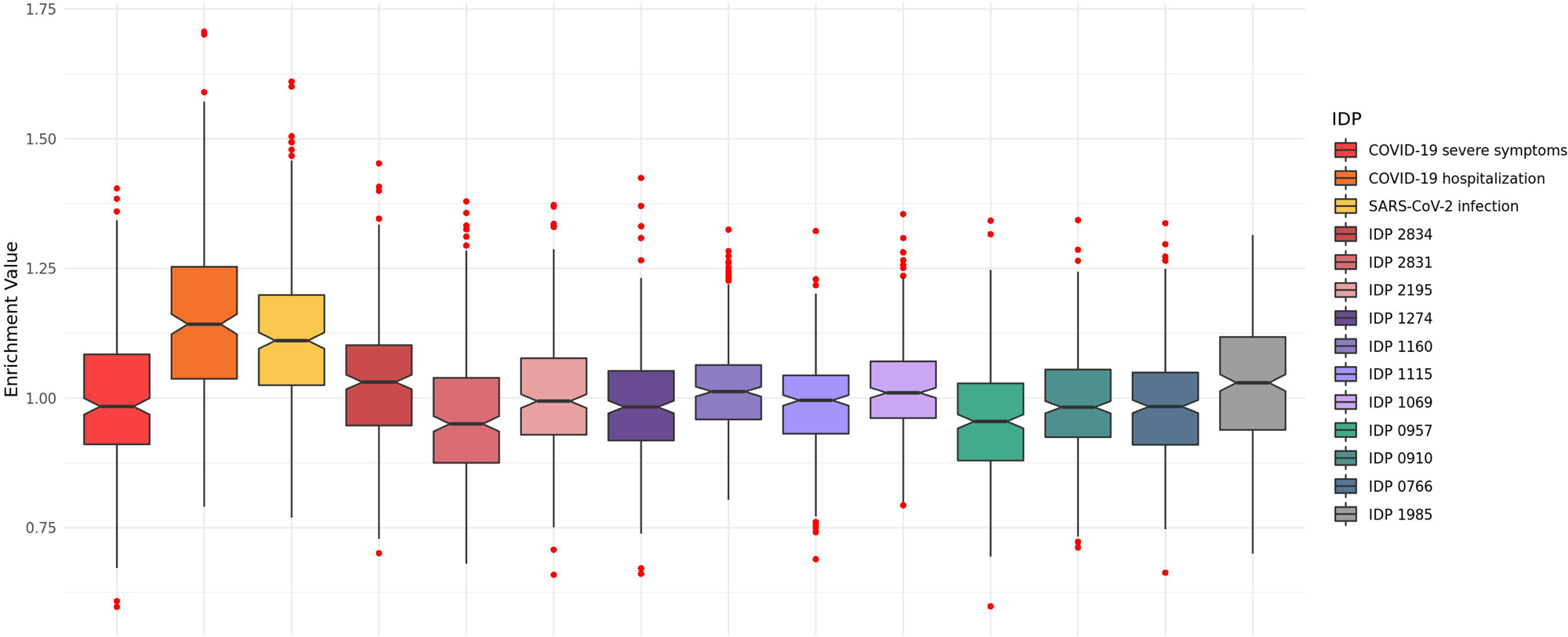
Brain imaging-derived phenotype (IDP) 2834 reflecting the edge 406 between nodes 28 (upper panel) and 29 (lower panel) of dimensionality 100 identified through spatial independent component analysis (ICA) of resting-state functional magnetic resonance imaging across three anatomical planes: axial (left), sagittal (top right), and coronal (bottom right)..

Considering the 11 brain IDPs identified by the global genetic correlation analysis, we uncovered five FDR-significant genomic regions with locus-specific pleiotropy related to COVID-19 hospitalization and SARS-CoV-2 infection (Table 1). Specifically, locus “3:181,424,324-183,203,155” showed genetic correlation of COVID-19 hospitalization with three brain IDPs related to parahippocampal and entorhinal cortical thickness (IDP 1069, 1115, 1160; rho ranging from 0.63 to 0.85). The IDPs related to parahippocampal cortical thickness (IDP 1069 rho=0.88; IDP 1160 rho=0.83) also showed local genetic correlation with COVID-19 hospitalization in locus “10:129,831,970-130,694,008”. In locus “22:49,559,208-50,347,996”, COVID-19 hospitalization was genetically correlated with two IDPs related to the cortical areas of the pars opercularis (IDPs 766 and 957; rho=-0.97). The sign of these local genetic correlations was in line with the ones observed in the global genetic correlation analysis (positive rg/rho with respect to cortical thickness and negative rg/rho with respect to cortical areas). Conversely, while rfMRI connectivity ICA100 edge 406 (IDP 2834) showed a positive global genetic correlation with SARS-CoV-2 infection, we identified “2:84,395,900-85,992,794” locus with local genetic correlation consistent with the one observed globally (rho=1) and “20:51,533,751-52,411,532” locus with inverse local genetic correlation (rho=-1).

**Table 1:**
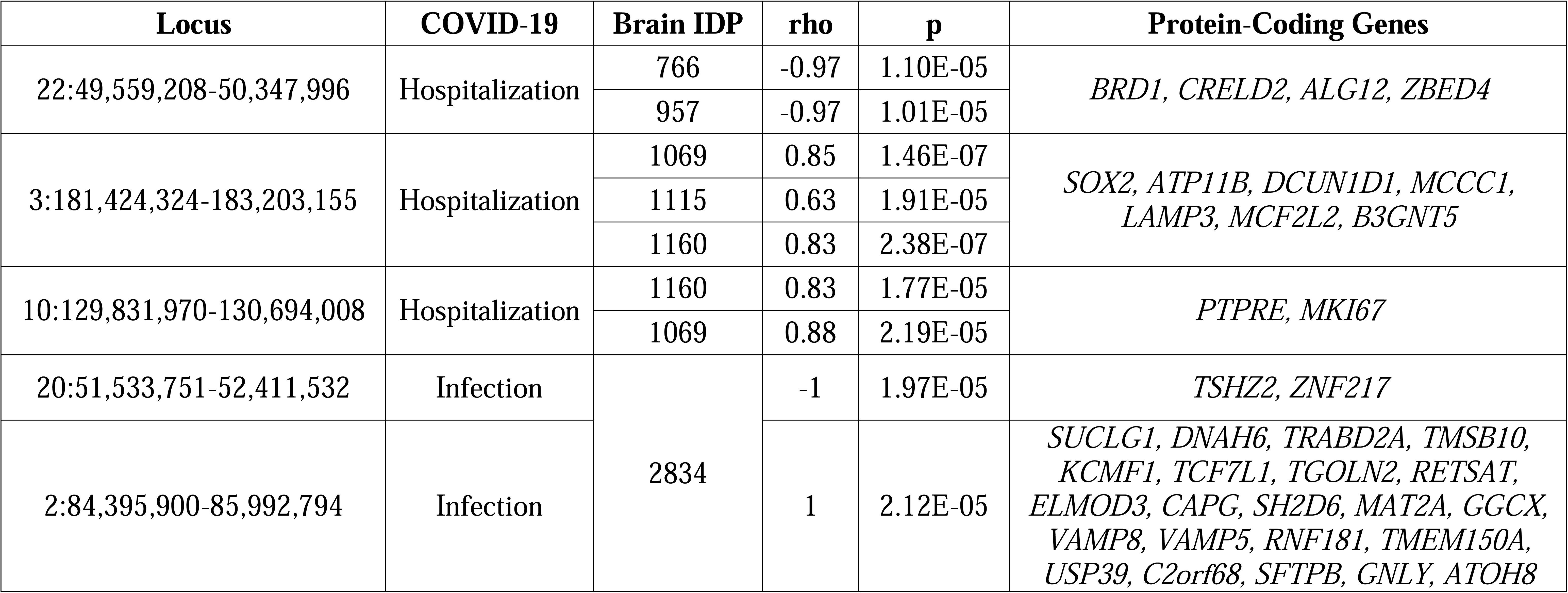
Local genetic correlations between COVID-19 outcomes and brain imaging-derived phenotypes (IDP) surviving false discovery rate multiple testing correction. IDP 766: Area of parsopercularis in the right hemisphere generated by parcellation of the pial surface using Desikan-Killiany parcellation; IDP 910: Area of Lat-Fis-ant-Horizont in the left hemisphere generated by parcellation of the white surface using Destrieux (a2009s) parcellation; IDP 957: Area of G-front-inf-Opercular in the right hemisphere generated by parcellation of the white surface using Destrieux (a2009s) parcellation; 1069: Mean thickness of parahippocampal in the right hemisphere generated by parcellation of the white surface using Desikan-Killiany parcellation; IDP 1115: Mean thickness of entorhinal in the right hemisphere generated by parcellation of the white surface using BA_exvivo parcellation; IDP 1160: Mean thickness of parahippocampal in the right hemisphere generated by parcellation of the white surface using DKT parcellation; IDP 1274: Mean thickness of G-oc-temp-med-Parahip in the right hemisphere generated by parcellation of the white surface using Destrieux (a2009s) parcellation; IDP 1985: Mean OD (orientation dispersion index) in medial lemniscus (right) on FA (fractional anisotropy) skeleton (from dMRI data); IDP 2195: rfMRI amplitudes (ICA100 node 32); IDP 2831: rfMRI connectivity (ICA100 edge 403); IDP 2834: rfMRI connectivity (ICA100 edge 406).

### Gene-Set and Drug-Repurposing Analyses

After Bonferroni multiple testing correction (p<6.31×10^-7^), we identified 305 GOs that were statistically enriched across the three COVID-19 outcomes tested and the 11 brain IDPs identified (Supplemental Table 3). Comparing GSA-MiXeR statistics, COVID-19 hospitalization and SARS-CoV-2 infection were more enriched for the shared GOs than severe COVID-19 respiratory symptoms and the 11 brain IDPs (Figure 3; Supplemental Table 3). The top shared GO enrichments suggested further biological convergence between COVID-19 and brain IDPs.

**Figure 3:**
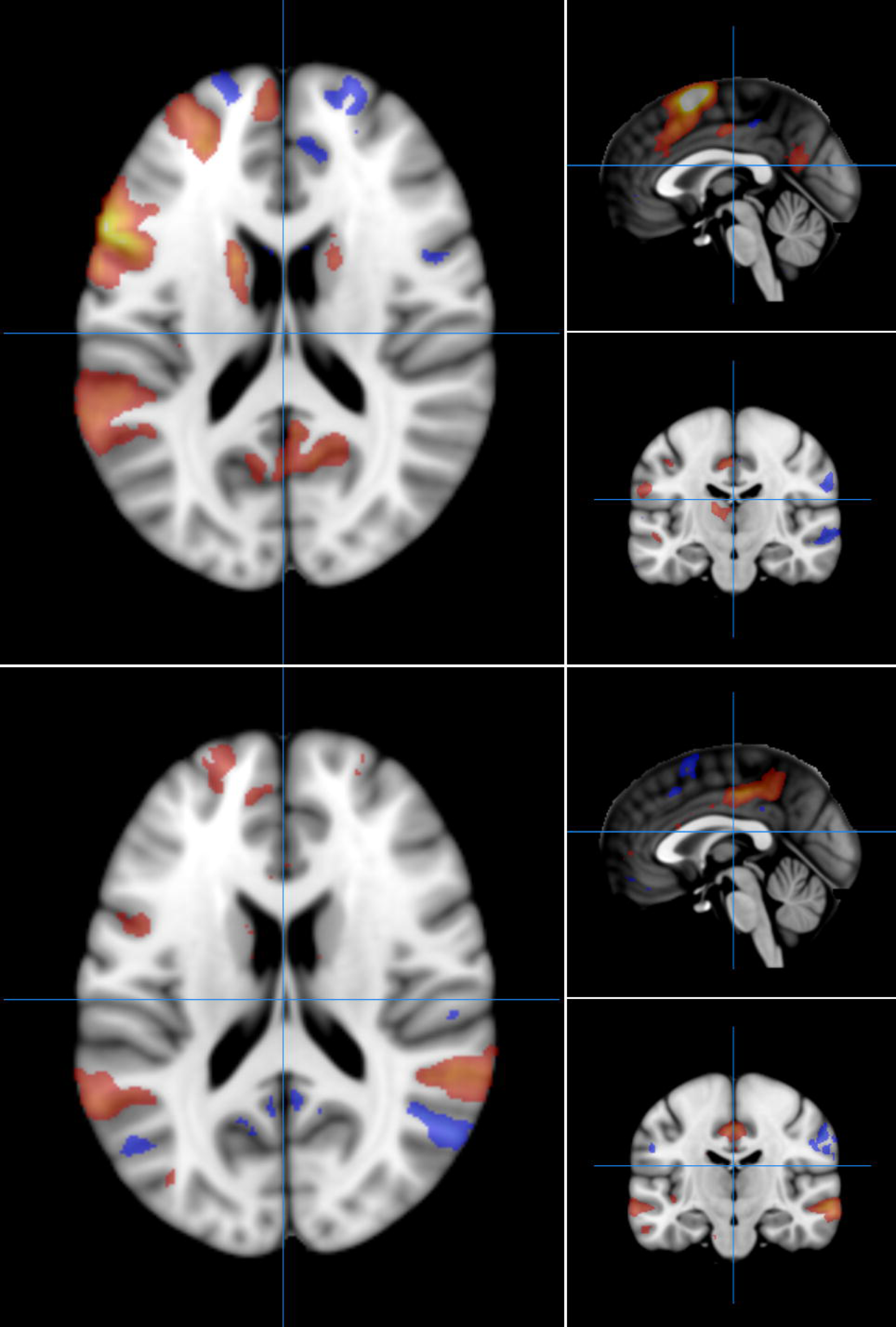
Enrichment distribution of the 305 Bonferroni-significant gene ontologies shared among COVID-19 outcomes investigated and the 11 brain imaging-derived phenotypes (IDP) identified. Full results are available in Supplemental Table 3.

For instance, tissue development (GO:0009888) was the most significant enrichment for COVID-19 hospitalization (enrichment=1.093, p=1.25×10^-23^), IDP 2831 (rfMRI connectivity ICA100 edge 403; enrichment=0.82, p=2.2×10^-62^), and IDP 2195 (rfMRI amplitudes ICA100 node 32; enrichment=0.981, p=1.5×10^-81^). Similarly, peptide biosynthetic process (GO:0043043) showed the highest enrichment for both SARS-CoV-2 infection (enrichment=1.61, p=9.39×10^-10^) and IDP 0766 (area of pars opercularis; enrichment=1.337, p=4.7×10^-15^). Interestingly, we also observed cross-domain enrichments. For instance, the highest enrichment of IDP 2834 (rfMRI connectivity ICA100 edge 406; enrichment=1.452, p=4.7×10^-17^) and IDP 1274 (mean thickness of the medial parahippocampal gyrus; enrichment=1.424, p=6×10^-21^) was with respect to innate immune response (GO:0045087). For SARS-CoV-2 infection, the most significant GO enrichment was synapse (GO:0045202; enrichment=0.912, p=1.44×10^-26^).

Through REVIGO approach, we identified 137 non-redundant Bonferroni-significant GOs related to biological processes (Supplemental Table 4). Of these, 109 were classified into six representative domains: cell projection organization (N=13), cellular response to stress (N=17), peptidyl-amino acid modification (N=20), regulation of intracellular signal transduction (N=30), tissue development (N=18), and vesicle-mediated transport (N=11). We observed neuroimmune convergence in both “tissue development” and “regulation of intracellular signal transduction” domains. Specifically, “tissue development” included GOs such as immune system development (GO:0002520), leukocyte differentiation (GO:0002521), lymphocyte activation (GO:0046649), neurogenesis (GO:0022008), nervous system process (GO:0050877), and head development (GO:0060322). “Regulation of intracellular signal transduction” domain included synaptic signaling (GO:0099536), regulation of immune system process (GO:0002682), and immune response (GO:0002764).

Considering FDR-significant GO enrichments shared across the three COVID-19 outcomes tested and the 11 brain IDPs identified, our drug-repurposing analysis identified 41 compounds (Supplemental Table 5). Fourteen of them were associated with downregulation of the pathways tested (enrichment score, ES<0). These included antiviral drugs (e.g., ribavirin ES=-0.519, p=4.15×10^-4^), heart medications (e.g., canadine ES=-0.504, p=6.68×10^-4^), chemotherapy medications (decitabine ES=-0.5, p=7.67×10^-4^), antihypertensive agents (epivincamine ES=- 0.468, p=4.25×10^-4^), antibiotics (e.g., ceforanide ES=-0.207, p=2.4×10^-4^), and antihistaminic medications (levocabastine ES=-0.195, p=6.47×10^-4^). While cardiovascular, anticancer, anti-infective, and antibacterial drugs were also among compounds associated with upregulated pathways (ES>0), these also included antipsychotic medications (clozapine ES=0.209, p=2.02×10^-4^; chlorprothixene ES=0.187, p=0.001) and antidepressants (nortriptyline ES=0.196, p=6.09×10^-4^).

## DISCUSSION

Integrating genome-wide information with multimodal brain MRI, our study provided new insights regarding pleiotropy among biological systems contributing to the relationships of COVID-19 outcomes with brain structure and function. Specifically, through the analysis of over 3,900 brain IDPs, we identified shared genetic effects linking SARS-CoV-2 infection and COVID-19 severe symptoms with brain activity, cortical thickness, and cortical areas.

With respect to rfMRI-generated IDPs, we observed different patterns in line with previous studies that reported that COVID-19 can alter brain activity, increasing or decreasing functional connectivity depending on the brain regions assessed ^29–31^. In our study, the connectivity network among Broca area, dorsolateral prefrontal cortex (dlPFC), anterior prefrontal cortex, posterior cingulate cortex, caudate, and left dorsal anterior cingulate cortex (IDP 2834, rfMRI connectivity ICA100 edge 406; Figure 2) and the brain activity related to the right supramarginal gyrus, primary somatosensory cortex, and motor cortex (IDP 2195, rfMRI amplitudes ICA100 node 32; Supplemental Figure 1) showed positive genetic correlation with COVID-19 outcomes.

Conversely, IDP 2831 (rfMRI connectivity ICA100 edge 403; Supplemental Figure 2) had an inverse genetic correlation with COVID-19 outcomes. This partially overlaps with IDP 2834, highlighting brain connectivity among anterior prefrontal cortex, caudate, Broca’s area, right dlPFC, and right dorsal anterior cingulate cortex. With respect to the brain regions underlying the rfMRI-generated IDPs identified in the present study, previous evidence supports their association with COVID-19 neurologic symptoms. For instance, dlPFC has been associated with cognitive deficits in young adults infected by SARS-CoV-2 ^32^. In comparison with healthy controls, patients with COVID-19 showed decreased regional homogeneity in the left angular gyrus, which was also negatively correlated with the severity of depression symptoms ^33^. Altered connectivity in the anterior cingulate gyrus and the left posterior cingulate gyrus has been reported in patients with post-COVID-19 syndrome ^30^. Additionally, tissue damage in the anterior cingulate cortex has been reported in mild COVID-19 cases ^34^, while the high expression of angiotensin-converting enzyme 2 (ACE2) in excitatory and inhibitory neurons and in non-neuron cells in the posterior cingulate cortex suggests the vulnerability of this brain region to pathogenetic processes of COVID-19 ^35^. With respect to Broca’s area, a reduction in the grey matter volume has been reported in COVID-19 patients ^36^. This may contribute to COVID-19 relationship with aphasia-like symptoms ^37^. Functional imaging studies have also linked COVID-19 outcomes to changes in the right superior temporal gyrus ^3^, the angular gyrus ^38^, the left caudate ^39^, and the supramarginal gyri ^38, 40^ , several of which overlap with the brain regions identified in our study.

With respect to cortical thickness, our findings highlight a positive genetic correlation with COVID-19 outcomes in parahippocampal and entorhinal gyri. Increased cortical thickness in the parahippocampal gyrus has been also reported i) in healthy COVID-19 survivors compared to non-infected controls and ii) in long-COVID-19 patients compared to healthy COVID-19 survivors ^41^. While our findings related to parahippocampal cortical thickness were specific to the right hemisphere, a previous study reported the association of the left parahippocampal region thickness reduction with cognitive dysfunction among patients affected by post-COVID-19 condition ^42^. Our findings related to the entorhinal cortex could be in line with the hypothesis that this region is linked to olfactory dysfunction and memory impairments observed in COVID-19 patients ^43^. Additionally, after intranasal inoculation in non-human primate models, SARS-CoV-2 RNA was detected in the entorhinal area and was associated with neuronal death, glial hyperplasia, and edema ^44^

In contrast to the positive genetic correlation observed with respect to cortical-thickness IDPs, COVID-19 outcomes showed an inverse genetic correlation with the cortical areas of the Lat-Fis-ant-Horizont area and the pars opercularis, which are involved in language production and syntax processing ^45^. Significant differences in the grey matter volume of the pars opercularis have been reported in acutely ill COVID-19 patients ^36^ and in COVID-19 survivors at one year after discharge ^46^. Additionally, decreased structural metrics of the pars opercularis were also associated with sleep disturbances in COVID-19 patients ^47^. With respect to anterior horizontal ramus of the lateral fissure (Lat-Fis-ant-Horizont), previous studies in critically ill COVID-19 patients or in individuals who died due to COVID-19 reported vascular alterations in this brain region ^48, 49^.

In addition to cortex-related IDPs, we also observed positive genetic correlations between COVID-19 and white matter tract orientation dispersion in the medial lemniscus, which is involved in sensory information processing and conscious proprioception ^50^. While no COVID-19 findings have been reported with respect to this specific brain region, altered orientation dispersion index of white matter across brain regions has been reported in post-COVID-19 aggravated or unchanged chronic insomnia ^51^.

The local genetic correlation analysis permitted us to identify five genomic regions (Table 1) that may play a key role in the pleiotropy linking COVID-19 to brain structural and functional variation. The local genetic correlations observed are likely due to the combined effects of multiple variants within each locus. Interestingly, we observed COVID-19 pleiotropy with different brain IDPs converged on the same loci. On locus “3:181,424,324-183,203,155”, COVID-19 hospitalization was genetically correlated with the three IDPs related to the cortical thickness of parahippocampal and entorhinal gyri (IDPs 1069, 1115, and 1160). Seven protein-coding genes map within this locus. Among them, *LAMP3* gene encodes an immune-related protein that was upregulated in hospitalized COVID-19 compared to non-hospitalized individuals ^52^. Another gene in this locus is *SOX2*, which plays an important role in brain development and cortical layer formation ^53^ and has been used to assess COVID-19 infection on the olfactory cell lineage ^54^. Also mapping within locus “3:181,424,324-183,203,155”, *ATP11B* and *B3GNT5* showed altered regulation during COVID-19 infection and variants in these genes have been associated with hippocampal synaptic plasticity impairment, cerebral small vessel disease, and glioblastoma ^55–57^, highlight their potential role in the development of COVID-19 related brain changes.

In locus “10: 129,831,970-130,694,008”, we also observed local genetic correlation between COVID-19 hospitalization and IDPs related to parahippocampal cortical thickness (IDPs 1160 and 1069). *MKI67* and *PTPRE* genes map in this region. The first encodes a protein involved in immune response to COVID-19 ^58^, while the second was observed to be upregulated during SARS-CoV-2 infection ^59^.

Locus “22:49,559,208-50,347,996” showed local genetic correlation between COVID-19 hospitalization and IDPs related to the cortical areas of the pars opercularis (IDPs 766 and 957). Among these genes mapping in this locus, *BRD1* has been implicated in immune reactivity and its downregulation was associated with increased B-cell infiltration and humoral immune response activation ^60^. Also located in this locus, *ALG12* protein product shares two pentapeptide sequences with SARS-CoV-2 spike glycoprotein and mutations in this gene have been linked to psychomotor retardation, immunodeficiency, hypotonia, and coagulation disorders ^61^.

We identified two chromosome regions (i.e., 2:84,395,900-85,992,794; 20:51,533,751-52,411,532) showing local genetic correlation between SARS-CoV-2 and IDP 2834 (rfMRI connectivity ICA100 edge 406; Figure 2). In locus “20:51,533,751-52,411,532”, *TSHZ2* has been implicated in brain development and it was one of the most significantly hypermethylated genes in COVID-19 patients six week after infection ^62^. In locus “2:84,395,900-85,992,794”, there are multiple genes involved in COVID-19 pathogenesis. *USP39* encodes an essential component for SARS-CoV-2 viral assembly and replication ^63^. *GGCX* is known to interact directly with viral proteins and has been suggested as potential target for COVID-19 drug development ^64^. *SFTPB*, *GNLY*, *ATOH8*, and *VAMP5* – also located in locus “2:84,395,900-85,992,794” – have been also linked with COVID-19 pathogenesis and outcomes ^65–68^. Together, these loci highlight potential molecular mechanisms and point out specific genes through which COVID-19 may exert pleiotropic effects on brain structure and function, mainly through pathways involved in immune response and neural integrity.

Moving from individual loci to biological processes, cellular components, and molecular functions, we identified 305 GOs that reached Bonferroni significance with respect to the three COVID-19 outcomes investigated and the 11 brain IDPs identified. The overall distribution of the enrichment statistics was largely consistent across these phenotypes (Figure 3; Supplemental Table 3), suggesting that the biological processes, cellular components, and molecular functions related to these GOs may have similar impacts on the three COVID-19 outcomes investigated and the 11 brain IDPs identified. In line with the expected systemic nature of the cross-domain pleiotropy linking COVID-19 to brain structure and function, several of the shared GOs were related to broad dynamics such as metabolic process, signal transduction, cellular morphogenesis, and ion transport. Some of these are expected to reflect pathogenetic processes that can affect both immune and central nervous systems. For example, mitochondrial dysfunction has been reported during COVID-19 acute and post-acute phases ^69^ and indicated as an important contributor to neural injury ^70^. In other cases, we identified enrichments that may reflect cross-domain relationships. In line with the role of innate immune response in the host defense against COVID-19 infections and the association of its dysregulation with neuroinflammation and neuronal damage in severe COVID-19 cases ^71^, we observed the highest enrichment for “innate immune response” GO (GO:0045087) for IDP 2834 (rfMRI connectivity ICA100 edge 406) and IDP 1274 (mean thickness of the medial parahippocampal gyrus). This supports an impact of innate immune response on brain structure and function potentially through its regulatory role of blood-brain barrier, microglial activation, and peripheral immune cell infiltration ^72^. Another interesting GO enrichment potentially linked to cross-domains relationships was “synapse” (GO:0045202), which was the most significant with respect to SARS-CoV-2 infection. Synapses are the fundamental units of communication between neurons and play a critical role in essential brain functions such as learning, memory, and cognition ^73^. Emerging evidence from studies highlighted how SARS-CoV-2 infection may disrupt synaptic function and integrity ^74, 75^. In this context, our findings support how pleiotropic mechanisms may contribute to COVID-19 neuropsychiatric comorbidities, pointing out specifically to those related to immune system, neurodevelopment and synaptic function.

Based on semantic similarities of the 305 shared Bonferroni-significant GOs, we identified 137 non-redundant terms and 109 of them were related to six domains reflecting broad biological dynamics. Among them, “tissue development” and “regulation of intracellular signal transduction” showed neuroimmune convergence. The GO term “tissue development” (GO:0009888) was the most significant for both COVID-19 hospitalization, IDP 2831, and IDP 2195 and other GO terms linked to this domain were related to the activation of immune system (i.e., immune system development, leukocyte differentiation, lymphocyte activation) and neurodevelopment (i.e., neurogenesis, nervous system process, head development). These findings appear to reflect the pleiotropy underlying crosstalk mechanisms between immune and central nervous systems that contribute to developmental processes ^76, 77^. This convergence may also extend to “regulation of intracellular signal transduction” domain, which included synaptic signaling (GO:0099536), regulation of immune system process (GO:0002682), and immune response (GO:0002764). An increasing number of studies reported various signaling mechanisms linking immune and central nervous systems, also highlighting their impact on health and disease ^78, 79^. Our results suggest that these interconnected pathways can also contribute to COVID-19 and its neuropsychiatric comorbidities.

Our drug-repurposing analyses identified 41 compounds with transcriptomic profiles matching the genes related to the GOs shared between COVID-19 outcomes and brain IDPs. In line with the systemic nature of the pleiotropic mechanisms linking COVID-19 to brain function and structure, we identified drugs used with respect to a range of clinical domains (e.g., cardiovascular diseases, cancer, and painful conditions) in addition to immune and neuropsychiatric conditions. Interestingly, while drugs related to other domains were associated with upregulation and downregulation of the pathways identified, all psychiatry-related compounds were associated with transcriptomic upregulation. These included antipsychotics (i.e., clozapine and chlorprothixene) and antidepressants (i.e., nortriptyline). Notably, while clozapine and nortriptyline have been associated with worse COVID-19 outcomes because of their immunosuppressive effect ^80, 81^, chlorpromazine appears to exert antiviral and immunomodulatory activity inhibiting the expression and secretion of IL-6 by monocytes activated by SARS-CoV-2 ^82^.

Our study has three key limitations that should be acknowledged. Due to the lack of large-scale GWAS in diverse populations, we investigated only data generated from populations of European descent. Therefore, our findings may not be generalizable to other population groups. Given the exploratory character of the present investigation, our initial assessment of the genetic correlation between COVID-19 outcomes and brain IDPs did not fully account for multiple testing correction. Finally, while our study successfully provided a valuable snapshot of the complex pleiotropic mechanisms linking COVID-19 outcomes to brain structure and function, further studies are needed to understand how shared biology contributes to the long-term neuropsychiatric effects of COVID-19.

In summary, this study provides novel insights into the genetic architecture shared between COVID-19 and brain structure and function. By leveraging large-scale genome-wide data and integrative analytical frameworks, we uncovered how neuroimmune pleiotropy affecting multiple biological processes could potentially contribute to COVID-19 impact on brain-related phenotypes. Overall, our findings strongly support the need to investigate COVID-19 and brain biology through studies assessing the interplay between central and peripheral systems.

## AUTHORSHIP CONTRIBUTION STATEMENT

Conceptualization: QC, RP; Methodology: JH, RP; Investigation: QC, JH, BCM, DQ, RP; Visualization: QC and JH; Funding acquisition: RP; Supervision: RP; Writing – original draft: QC, RP; Writing – review & editing: QC, JH, BCM, DQ, RP.

## DATA AVAILABILITY

COVID-19 genome-wide association statistics are available at https://www.covid19hg.org/results/r7/. Genome-wide association statistics of brain imaging-derived phenotypes are available at https://open.win.ox.ac.uk/ukbiobank/big40/. The data generated from the present study are included in the manuscript and its supplemental material.

## Data Availability

The data generated from the present study are included in the manuscript and its supplemental material.

## ACKNOWLEDGMENTS

This study was supported by a grant from the National Institutes of Mental Health (RF1 MH132337). JH also acknowledges support from the American Foundation for Suicide Prevention (PDF-0-065-23). The authors thank the COVID-19 Host Genetics Initiative and the Oxford Brain Imaging Genetics Study for making their data available.

