## Supplemental Figures for "Neuroimmune pleiotropy links COVID-19 outcomes to brain structural and functional imaging-derived phenotypes"

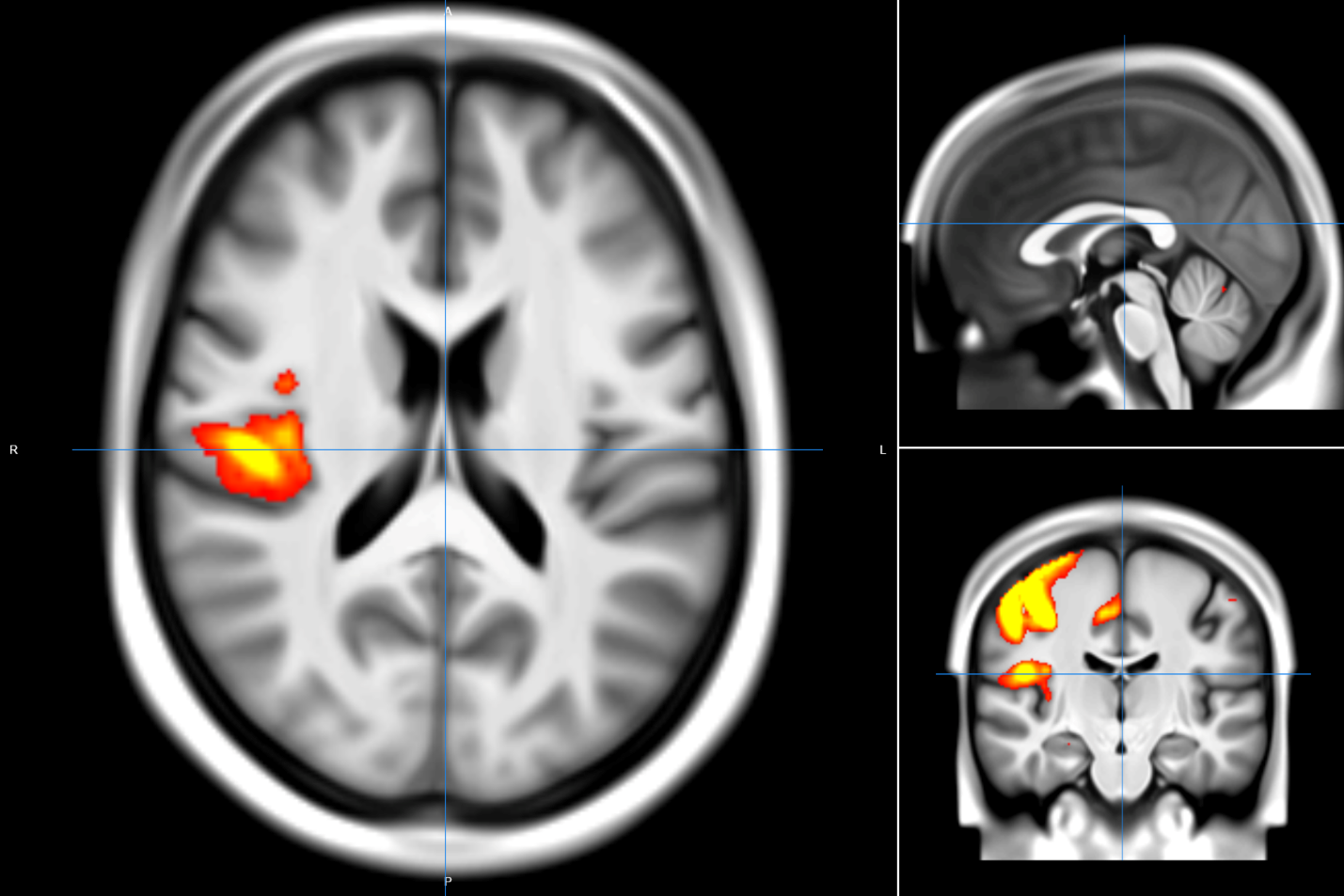

**Supplemental Figure 1**: Brain imaging-derived phenotype (IDP) 2195 reflecting the amplitudes of node 32 of dimensionality 100 identified by spatial Independent Component Analysis (ICA) of resting-state functional magnetic resonance imaging across three anatomical planes: axial (left), sagittal (top right), and coronal (bottom right).

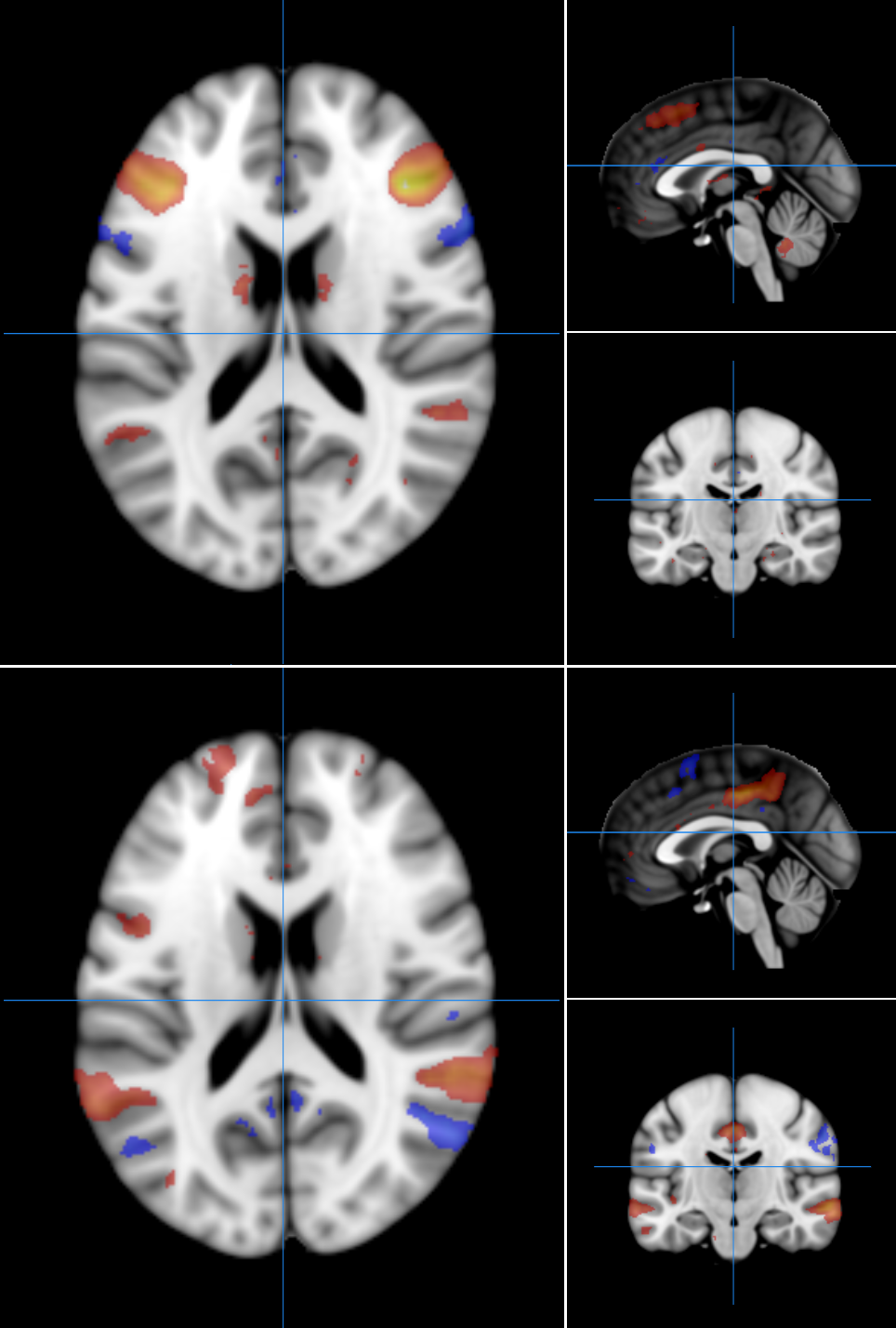

**Supplemental Figure 2**: Brain imaging-derived phenotype (IDP) 2831 reflecting the edge 403 between nodes 25 (upper panel) and 29 (lower panel) of dimensionality 100 identified by spatial Independent Component Analysis (ICA) of resting-state functional magnetic resonance imaging across three anatomical planes: axial (left), sagittal (top right of each panel), and coronal (bottom right of each panel).
